# Insurance Churn and Survival After Heart Transplantation in the Modern Era: A National Cohort Study

**DOI:** 10.1101/2025.10.01.25337134

**Authors:** Ye In Christopher Kwon, David T. Zhu, Andrew Min-Gi Park, Michael Keller, Matthew Ambrosio, Jay Patel, Motaz Al-Yafi, Vigneshwar Kasirajan, Zubair A. Hashmi

## Abstract

**Background:** Heart transplantation (HT) in the United States continues to be increasingly performed in the setting of expanding Medicare and Medicaid public insurance coverage. We analyze the impact of recipient insurance trajectories on long-term graft and recipient survival following the 2018 United Network for Organ Sharing (UNOS) heart allocation change.

**Methods:** Adults aged 18–64 undergoing first-time HT between 10/2018 and 3/2024 were identified in the UNOS database. Patients surviving ≥1 year were stratified by insurance trajectory (private vs. public, including Medicare, Medicaid, and VA) at waitlist, transplantation, and 1-year follow-up. Kaplan-Meier and Cox regression models were used to assess survival and graft failure risk.

**Results:** Among 15,864 patients, 42.5% had continuous private insurance, 33.9% had continuous public insurance, 12.6% transitioned from private to public, 4.8% transitioned from public to private, and 6.3% experienced multiple transitions. Five-year survival (p<0.0001) and graft survival (p=0.03) were poorer in patients with continuous public insurance or who transitioned from private to public insurance. Primary graft dysfunction rates were highest among recipients with continuous public insurance (1.4%; p<0.0001). Continuous public insurance (HR 1.27, p<0.0001) and multiple transitions (HR 1.19, p=0.04) increased mortality risk compared to continuous private insurance. Continuous public insurance (HR 1.29, p=0.01), private-to-public transition (HR 1.32, p=0.03), and multiple transitions (HR 1.09, p=0.02) were linked to higher graft failure risk. Public-to-private transition was associated with lower graft failure risk (HR 0.73, p=0.02). Residence in distressed communities increased mortality (HR 1.21, p=0.003) and graft failure risk (HR 1.31, p=0.001).

**Conclusions:** Public insurance and insurance instability are associated with worse HT outcomes, while gaining private insurance was associated with improved outcomes, highlighting disparities in post-transplant care.

**What is Known:**

1. Post-heart-transplant outcomes differ by insurance type, with publicly insured recipients generally experiencing worse long-term survival and higher complication rates than privately insured recipients.
2. Prior transplant analyses often treat insurance as a static exposure rather than a time-varying trajectory spanning waitlist, transplant, and follow-up.
3. Insurance transitions (“churn”) are detrimental: in pre-Affordable Care Act and the 2018 allocation change cohorts, private to public switch within 1-year associates with higher mortality.
4. Socioeconomic disadvantage independently correlates with poorer post-transplant outcomes beyond clinical risk.

**What the Study Adds:**

1. This is the first national, post–2018 allocation analysis to model insurance as a time-varying trajectory across the waitlist, transplant hospitalization, and the first post-transplant year.
2. Continuous public insurance and insurance instability (multiple coverage transitions) are each independently associated with higher adjusted risks of mortality and graft failure than continuous private insurance.
3. The direction of churn is clinically meaningful: a private to public switch increases graft-failure risk, whereas public to private associates with lower mortality and graft failure rates compared with remaining on public insurance
4. Defines a distinct and actionable morbidity profile linked to continuous public coverage—longer hospitalizations and higher rates of dialysis, acute rejection, and treatment for rejection, along with the highest incidence of primary graft dysfunction — pinpointing concrete targets for perioperative pathways, discharge planning, and payer-continuity interventions.
5. Neighborhood socioeconomic distress independently predicts mortality and graft failure above and beyond clinical covariates and insurance trajectory, highlighting structural levers for quality-improvement and policy.

## Introduction

Access to heart transplantation (HT) in the United States has mainly been restricted to patients with sufficient insurance coverage, as the ability to afford transplantation and lifelong post-operative care is a key factor for transplant eligibility. Historically, most HT recipients have been covered by private insurance or Medicare. For instance, in a national cohort from 2002 to 2011, only 12.5% of HT recipients had Medicaid, compared to 57.6% with private insurance and 29.9% with Medicare. However, these trends have changed significantly. Public programs now fund nearly half of all transplants, with their share rising from 28.2% in 1997 to 48.8% in 2016, driven by Medicaid growth (9.4% to 15.5%) and Medicare expansion (18.2% to 30.3%). This increase probably reflects policy changes such as Medicaid expansion under the Affordable Care Act (ACA) and the growing number of older or disabled patients qualifying for coverage. Notably, insurance coverage patterns also differ based on patient demographics: Medicaid-funded HT patients tend to be younger, female, non-White, and have lower socioeconomic status.

Insurance churn — changes in insurance coverage type over time — has emerged as an important yet understudied factor in transplant outcomes. Churn can occur when a patient loses employer-sponsored insurance due to declining health and transitions to Medicaid or Medicare or moves from public to private coverage through new employment or spousal insurance^6^. Such changes could disrupt continuity of care, medication access, and relationships with providers, potentially affecting outcomes^6^. One study in heart failure patients found that churn was associated with a 16% longer length of stay, 32% higher in-hospital mortality, and 18% greater hospital costs, likely from coverage gaps and interrupted care^7^. However, this study did not examine the trajectory of churn (e.g., private to public, or vice versa)^7^. To our knowledge, only one peer-reviewed study has evaluated insurance trajectory patterns in HT recipients. Tumin et al. analyzed a 2006–2013 cohort (pre-ACA period) and found that patients who switched from private to public insurance within 1-year post-transplant faced a 25% higher mortality hazard compared to those with continuous private insurance^8^. Patients who moved from public to private insurance had a 22% lower mortality hazard than those remaining on public insurance^8^.

Notable gaps remain in the literature. Most prior analyses are based on cohorts from the early 2000s to early 2010s, before major changes such as the ACA’s 2014 implementation and the 2018 heart allocation policy revision-which could impact both insurance coverage patterns and transplant outcomes. Additionally, prior studies have treated insurance type as a fixed variable, limiting insight into how time-varying patterns of insurance churn affect post-transplant outcomes. Finally, most work has largely focused on mortality, with less attention to other complications. Our study aims to address these gaps by examining multiple insurance trajectories, including repeated coverage changes, in recent years and across a broader set of outcomes. These findings may help guide targeted interventions and policies to improve equity and outcomes for HT patients affected by insurance churn.

## Materials and Methods

### Data source

De-identified, patient-level data were obtained from the nationwide Organ Procurement Transplantation Network (OPTN), which is facilitated by the United Network for Organ sharing (UNOS). This study qualified for exemption from review by the Virginia Commonwealth University Institutional Review Board. This study adheres to the Strengthening the Reporting of Observational Studies in Epidemiology reporting guidelines for cohort studies.

### Study population

The Standard Transplant Analysis and Research (STAR) files were queried to identify adult patients (≥18 years) listed for first-time HT in the United States from 10/18/2018 to 3/31/2024 (all post-2018 heart allocation policy changes and post-Affordable Care Act). Exclusion criteria included pediatric patients, multi-organ transplants, and those who did not survive for at least 1 year (**Figure 1**). Recipients were stratified by insurance status at waitlisting, transplantation, and 1-year follow-up. Insurance status was categorized as private (including self-pay), public (Medicare, Medicaid, or Veterans Affairs [VA]), or other (e.g., foreign government, free/donation-based care). Following Tumin et al.’s methodology^8^, we categorized insurance churn into seven trajectories: (1) continuous private; (2) continuous public; (3) transition from private to public; (4) transition from public to private; (5) multiple insurance transitions; (6) any trajectories involving other insurance types; and (7) any trajectories with incomplete data.

**Figure 1.**
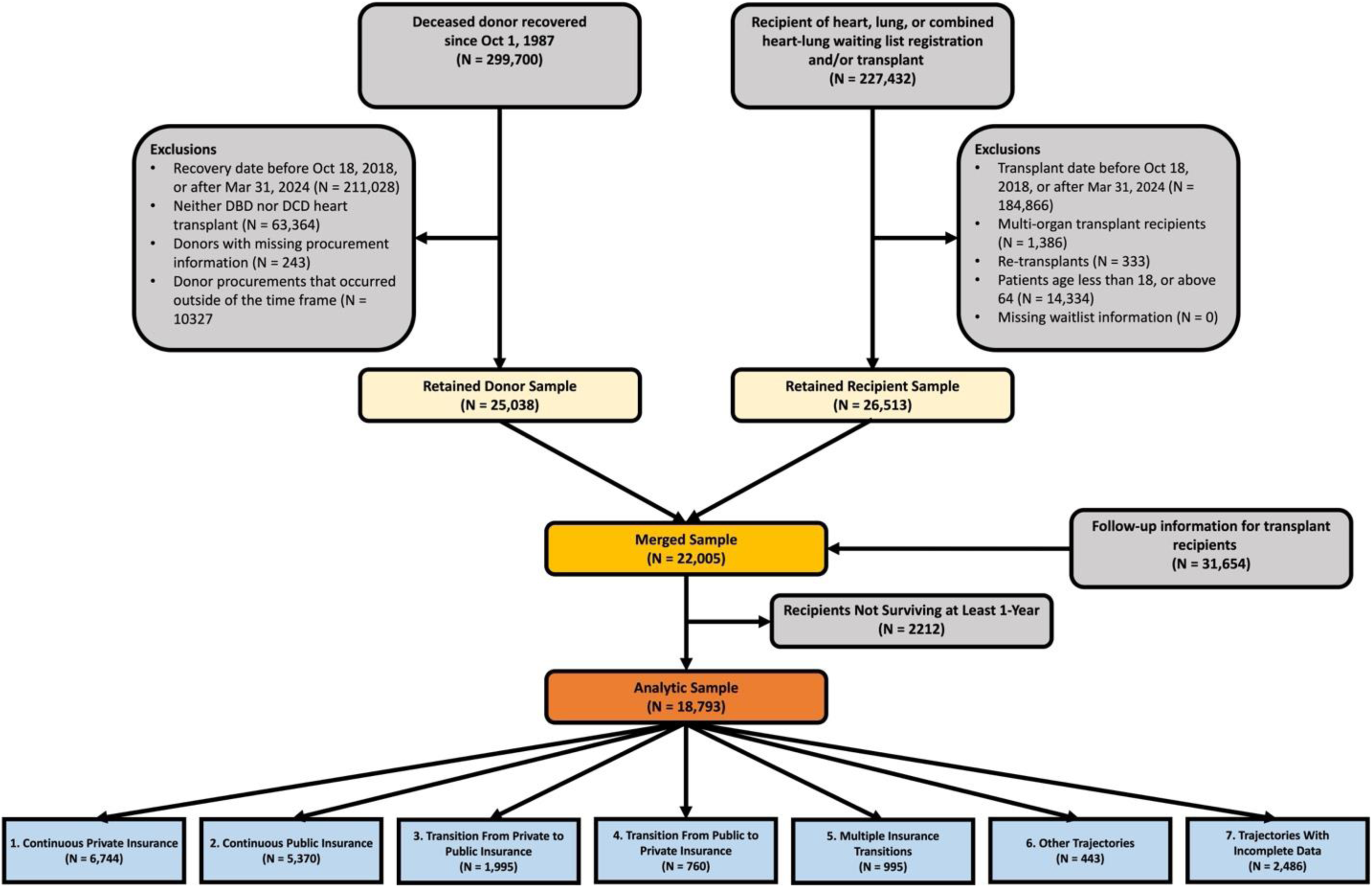
Flow chart for patient inclusion and exclusion using the United Network for Organ Sharing registry.

Trajectories (3) and (4) reflected a single change in insurance type—either at the time of transplant or at the first-year follow-up—while trajectory (5) represented patients who switched insurance types at transplant but later reverted to their original coverage during follow-up.

### Outcomes and definitions

The primary outcome was recipient survival at 1-, 3-, and 5-years after HT, with follow-up data available through June 2025. We also examined the rates of acute rejection, permanent pacemaker implantation, stroke, dialysis, treatment for rejection within 1-year, and post-HT length of hospital stay (LOS). Graft failure was based on the definition by the International Society for Heart & Lung Transplantation’s (ISHLT)’s 32^nd^ Adult Heart Transplant Report^9^. For donation after circulatory death (DCD) recipients, normothermic regional perfusion (NRP) was distinguished from direct procurement and preservation (DPP) according to previously described methodologies^10–12^. To account for neighborhood-level socioeconomic disadvantage, we calculated the Distressed Communities Index (DCI), a composite score derived from seven weighted indicators of community distress. Residence ZIP codes in the highest quintile of DCI values were classified as distressed^13^.

### Statistical analysis

Baseline donor and recipient characteristics were compared between groups using χ² and Fisher’s exact test for categorical variables and Welch’s two-sample t-test for continuous variables. Continuous variables are reported as means with standard deviations (SD), and categorical variables as numbers with percentages. Unadjusted recipient and graft survival was assessed using Kaplan-Meier time-to-event analyses, and risk-adjusted outcomes were evaluated using multivariate Cox proportional hazard models with hazard ratios (HRs) reported with 95% confidence intervals (CI). Model covariates were selected using the least absolute shrinkage and selection operator to enhance model discrimination and minimize collinearity. Additional assessment of collinearity was performed using variance inflation factors (VIF) and pairwise correlation analysis. The average VIF was 1.27, and all VIF values were below 5, indicating no significant multicollinearity.

Covariates were selected based on biological plausibility and confounding factors. This includes demographic (recipient and donor age, sex, body mass index [BMI], race/ethnicity, residence in distressed communities, and donor-recipient distance) and clinical (comorbidities, preoperative labs, hemodynamics, pre-HT circulatory support, DCD status, procurement method, *ex-vivo* machine perfusion, and allograft ischemia time) factors. Insurance churn trajectories were modeled as piecewise-constant time-varying covariates, coded as 0 before and 1 after the earliest post-transplant transition. Missing data were handled by multiple imputations. All statistical analyses were conducted using SAS (version 9.4; SAS Inc., Cary, NC, USA). All *p*-values were based on two-sided statistical tests, with significance at *p*<0.05.

## Results

### Insurance Trajectories Post-HT

We included 22,005 recipients of HT during the study period (**Figure 1**). In the main analysis, 2,212 cases were excluded due to mortality or censoring within the first year of transplant. For comparison of descriptive statistics and multivariable analyses, an additional 2,929 cases were excluded if they had incomplete coverage data or their mode of payment was other than private, self-pay, Medicaid, Medicare, or the VA. Thus, primary analyses were performed on 15,864 HT recipients. Most common trajectories were continuous private (42.5%, N=6,744), followed by continuous public (33.9%, N=5,370), transition from private to public (12.6%, N=1,995), multiple transitions (6.3%, N=995), and transition from public to private (4.8%, N=760), and. By the time of transplantation, insurance status showed significant transitions, with 64% of private-to-public transition recipients maintaining private coverage and 36% switching to public coverage, while 66.6% of public-to-private recipients remained on public insurance (**Table 1**). Temporal trends from 2018 through 2023 revealed increasing volumes of HT across all insurance categories (**Figure 2**). Notably, continuous public insurance (*β*=45.2; p<0.0001) and multiple insurance transitions (*β*=15.3; p<0.0001) demonstrated statistically significant increases. Continuous private insurance maintained the highest overall volume despite relatively stable numbers throughout the study period.

**Figure 2.**
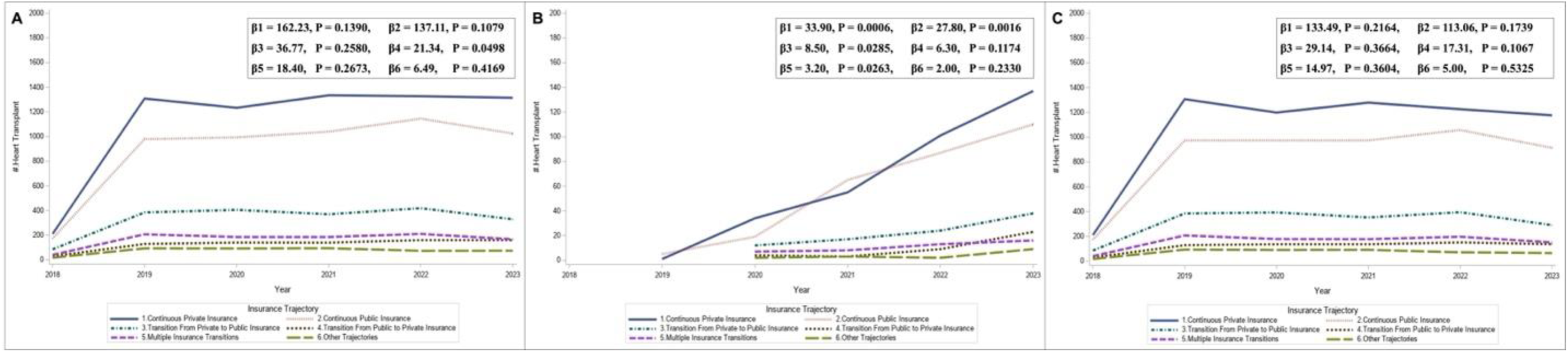
Temporal trends in overall (**A**), DCD (**B**), and DBD (**C**) HT by recipient insurance trajectory in the United States, 2018 – 2023. Slope estimates (β) and P values from linear regression are shown for each group.

**Table 1.**
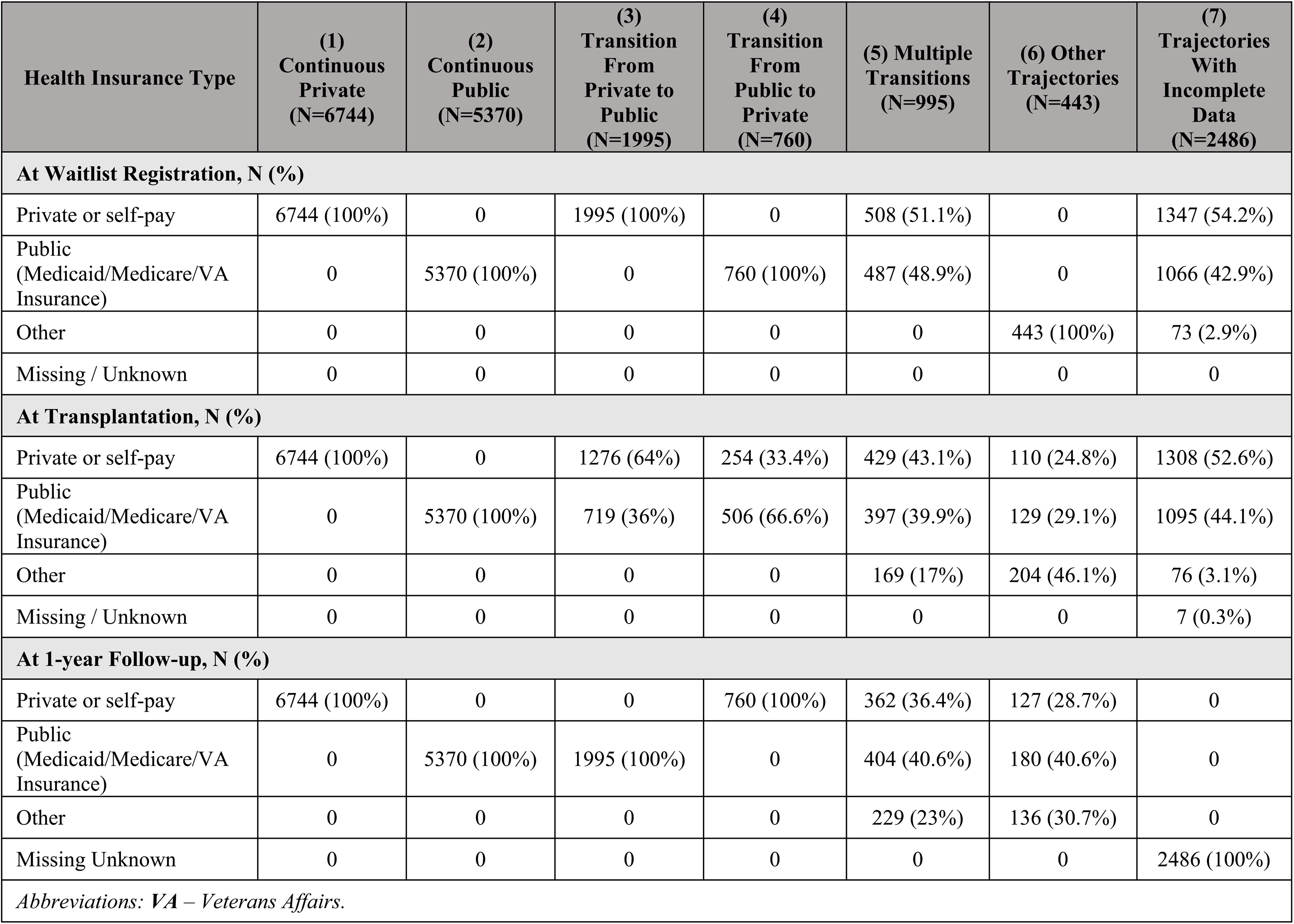
Types of recipients’ health insurance at waitlisting, transplantation, and 1-year follow-up by insurance trajectories.

### Baseline Characteristics

Significant demographic differences were observed across these groups (**Table 2**). Black recipients were most prevalent in the continuous public group (31%; *p*<0.0001), while Hispanic/Latino recipients were highest among multiple transitions (14.6%; *p*<0.0001).

**Table 2.**
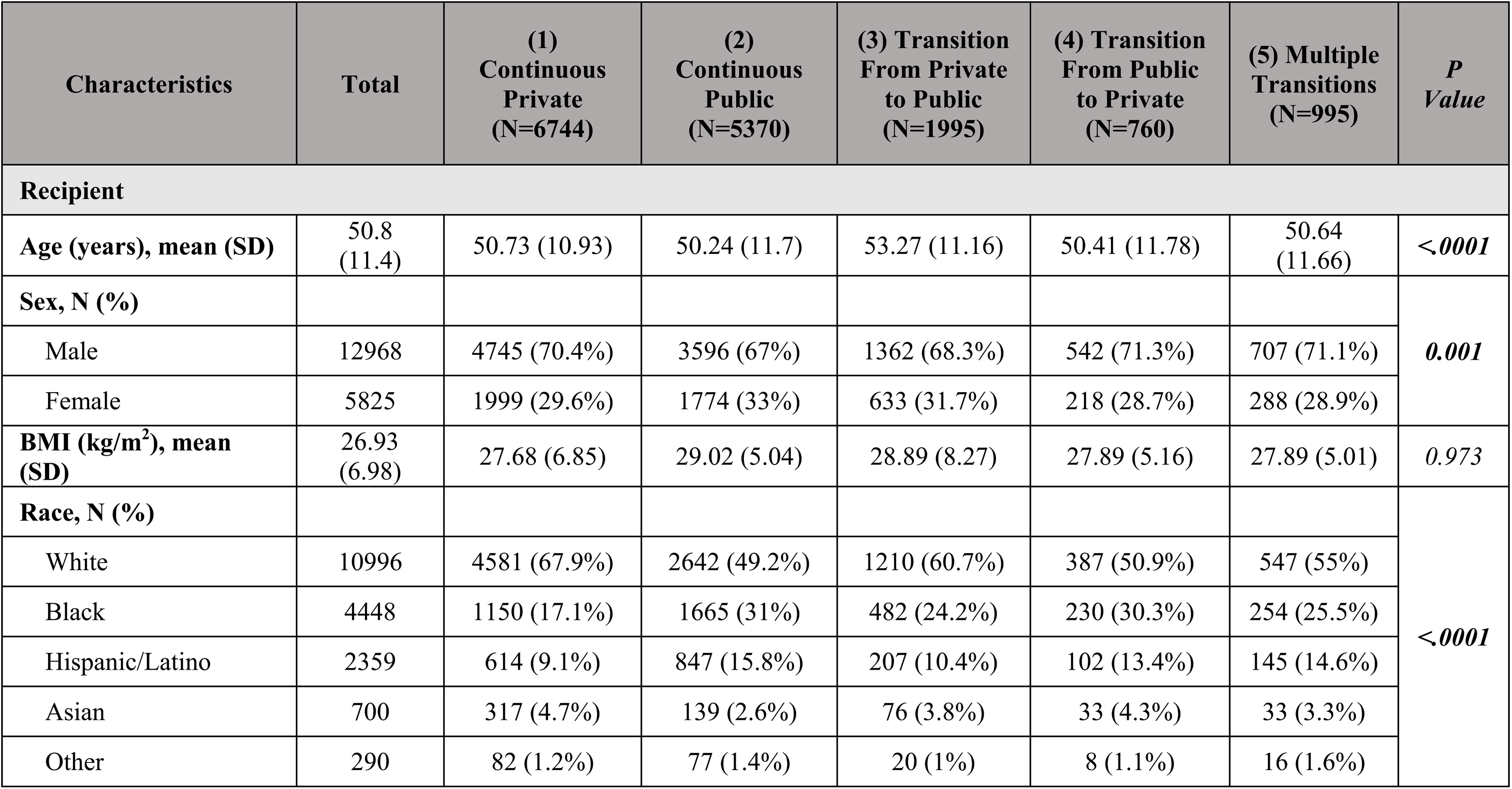
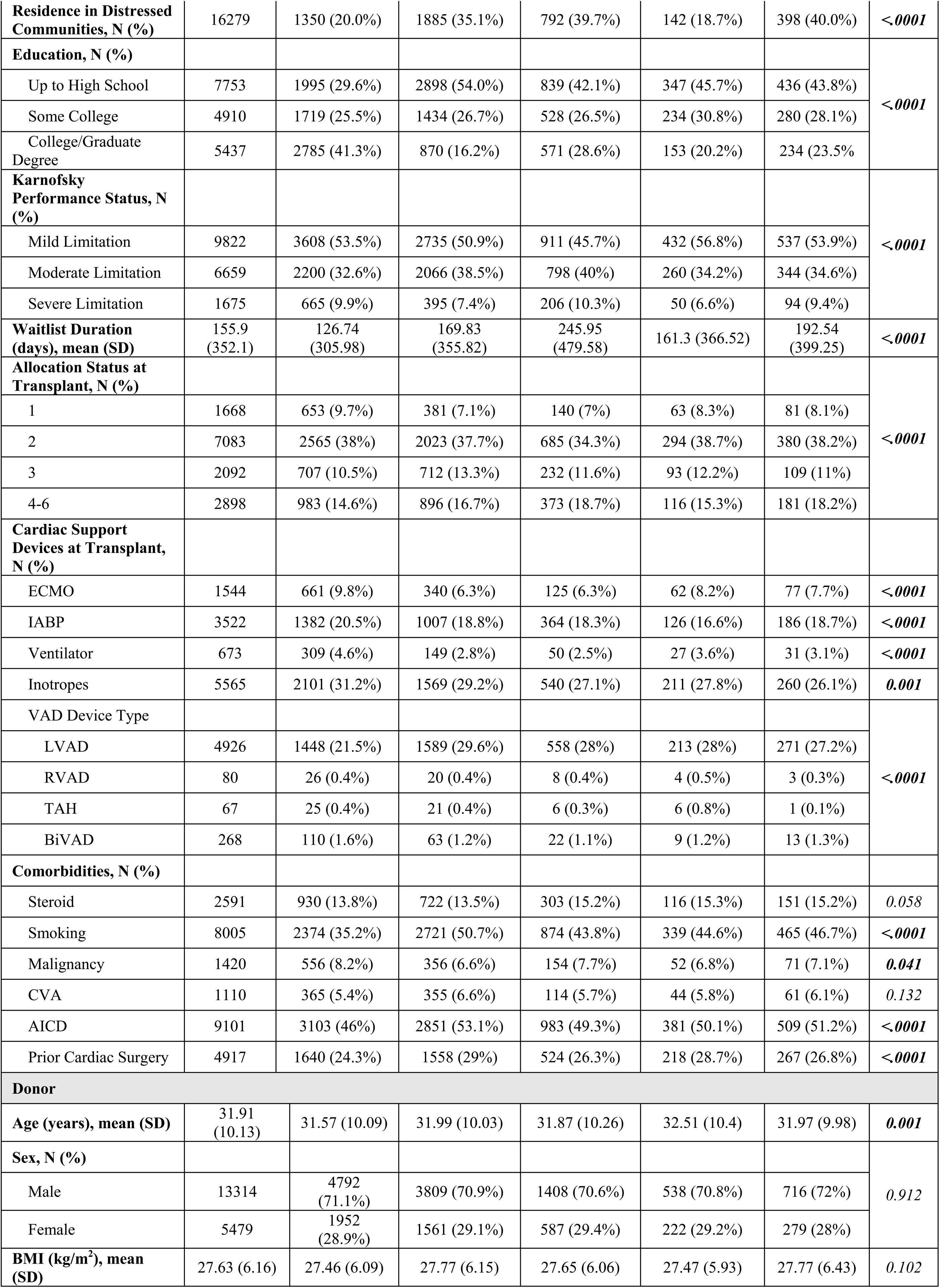
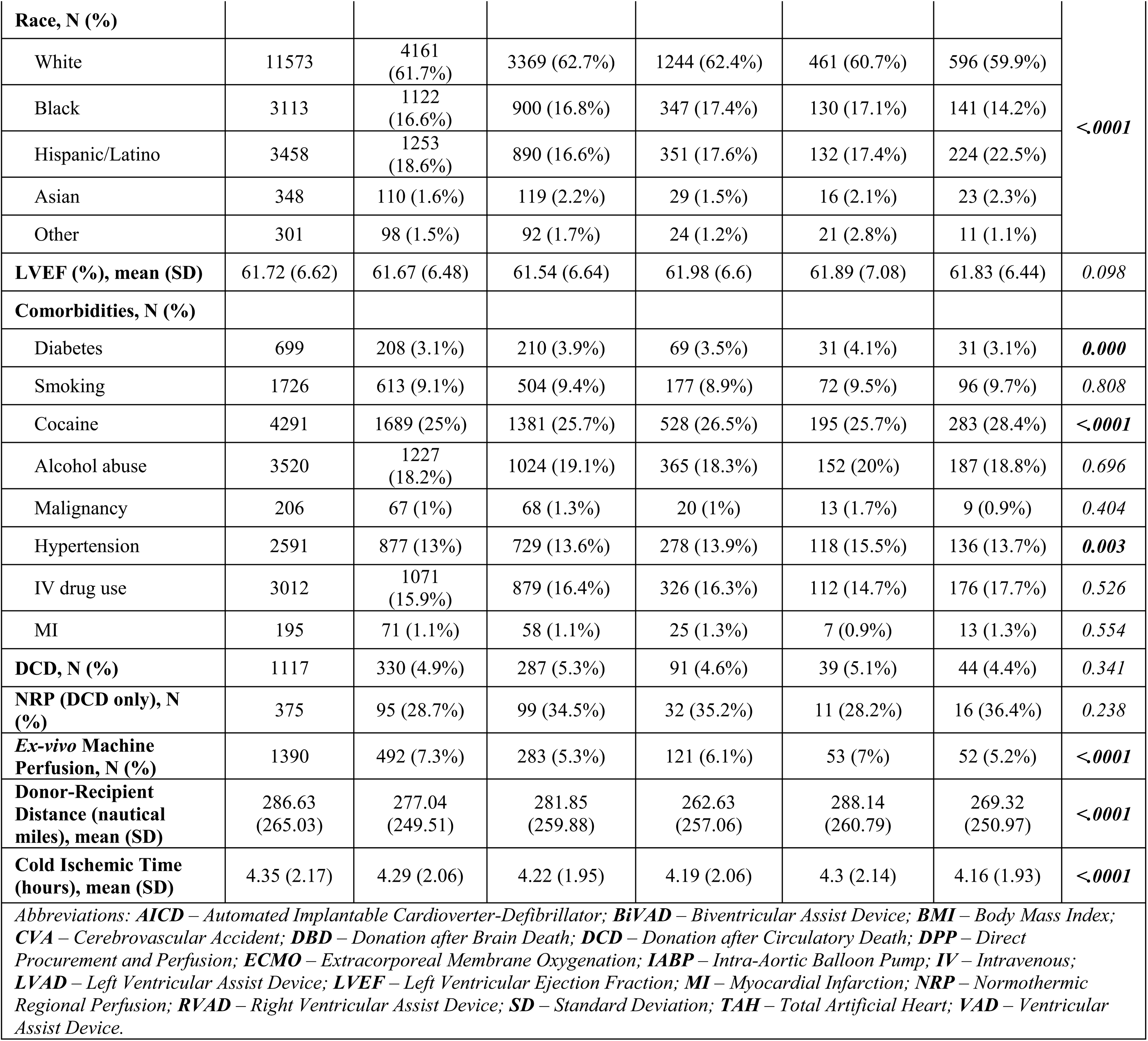
Baseline recipient and donor characteristics by insurance trajectories.

Recipients with continuous public insurance and multiple insurance transitions were significantly more likely to reside in distressed communities (continuous public: 35.1%, multiple transitions: 40.0%; *p*<0.0001). They also had lower levels of educational attainment compared to recipients with continuous private insurance (*p*<0.0001). Furthermore, the continuous public and multiple transitions groups exhibited higher prevalence of smoking (continuous public: 50.7%, multiple transitions: 46.7%; *p*<0.0001) and were less likely to receive ECMO (continuous public: 6.3%; *p*<0.0001). There were no variations in DCD (*p*=0.341) or NRP (*p=*0.238) usage across groups. However, *ex-vivo* machine perfusion was most common among recipients with continuous private insurance (7.3%) followed by public-to-private transitions (7%; *p*<0.0001).

### Clinical Outcomes

Insurance trajectories had significant impacts on clinical outcomes post-HT (**Figure 3**). Unadjusted overall mortality was highest among continuous public insurance recipients (15.5%) and those experiencing multiple transitions (14.9%) compared to continuous private insurance (12.1%; *p*<0.0001). Mortality at 1-, 3-, and 5-years consistently showed significantly higher rates for continuous public recipients (4.6%, 12.4%, and 15.1%, respectively) and multiple transition recipients (4.7%, 12%, and 14.6%, respectively) compared to continuous private recipients (4.5%, 9.9%, and 11.9%, respectively) (**Table 3**). Accordingly, multivariable Cox proportional hazards models confirmed increased mortality risk for recipients with continuous public insurance (HR 1.27, 95% CI 1.15-1.40; *p*<0.0001) and multiple insurance transitions (HR 1.19, 95% CI 1.12-1.43; *p*=0.044), compared to continuous private insurance (**Table S1**).

**Figure 3.**
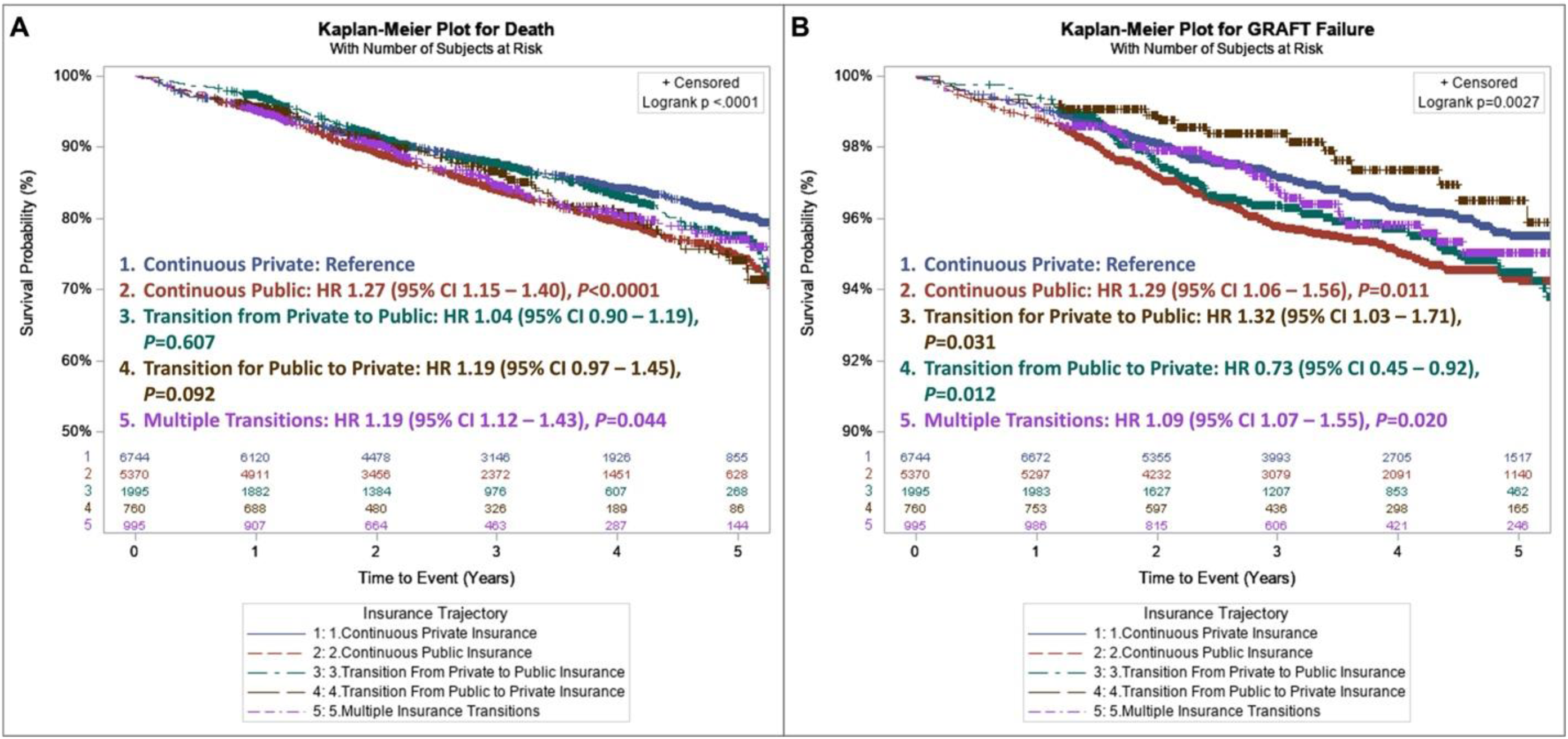
Kaplan-Meier recipient (**A**) and graft (**B**) survival curves and numbers of patients at risk by known insurance trajectories between waitlisting and 1-year after HT. Hazard ratios (HR) with 95% confidence intervals (CI) are from multivariable Cox models. Log-rank P values compare trajectories.

**Table 3.**
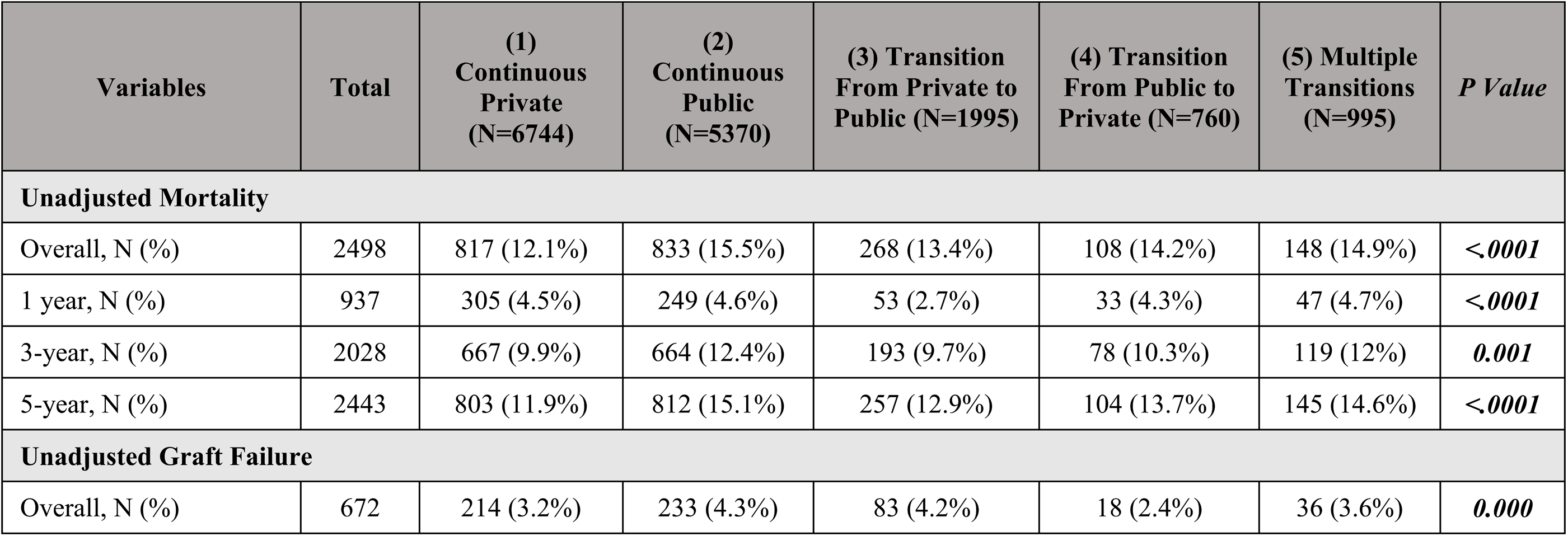
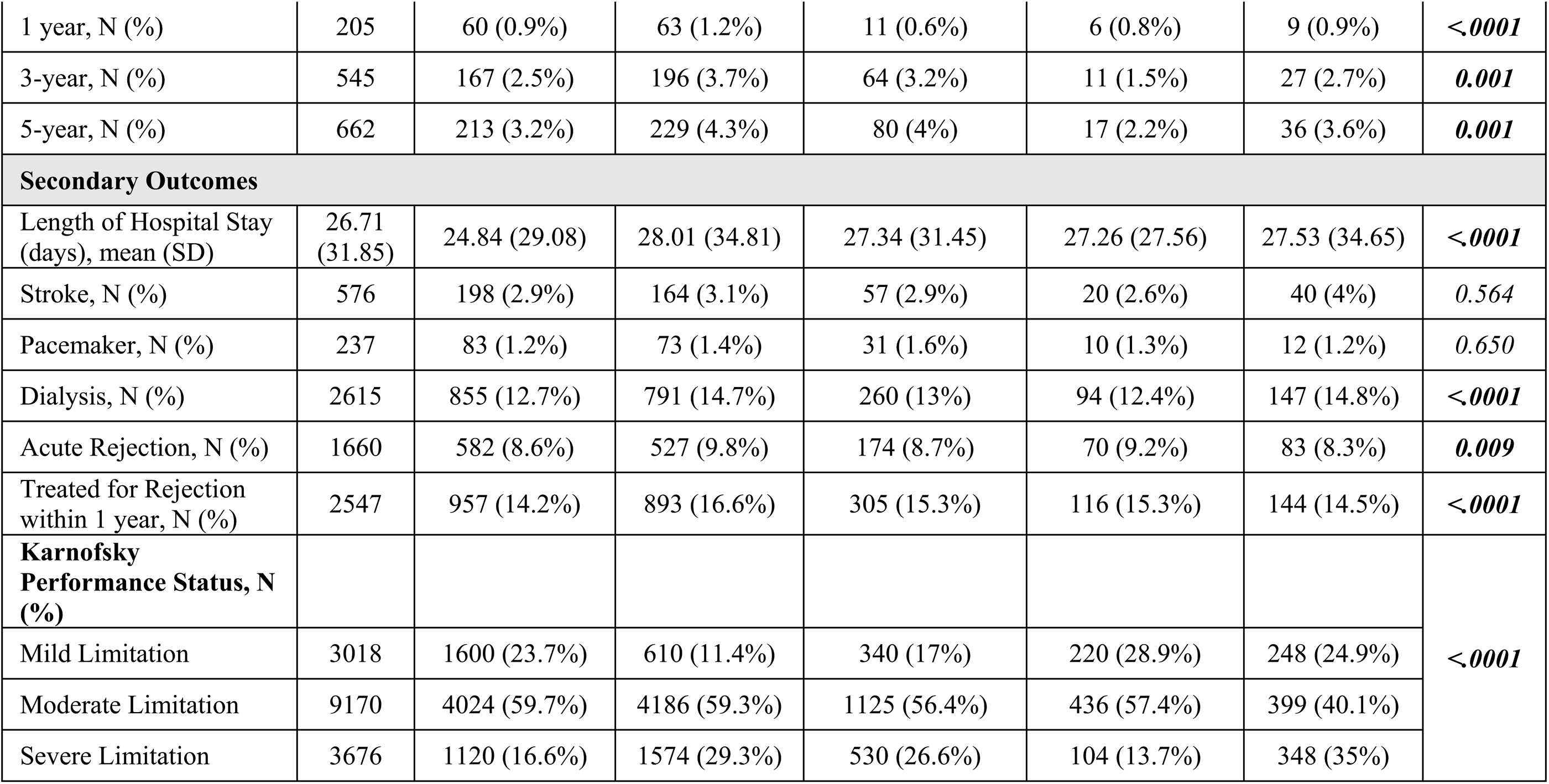
Post-HT outcomes by insurance trajectories.

Transitioning from private to public insurance did not significantly increase mortality risk (HR 1.04, 95% CI 0.90-1.19; *p*=0.607). Importantly, residence in distressed communities was independently associated with increased mortality risk (HR 1.21, 95% CI 1.12-1.57; *p*=0.003), whereas level of educational attainment beyond college degrees was associated with decreased mortality risk (HR 0.51, 95% CI 0.32-0.80; *p*=0.004).

Unadjusted graft failure rates followed a similar pattern, with overall graft failure highest in continuous public (4.3%) and private-to-public transition groups (4.2%; *p*=0.001). At 1-, 3-, and 5-years post-HT, continuous public insurance recipients consistently demonstrated higher graft failure rates (1.2%, 3.7%, and 4.3%, respectively) compared to continuous private insurance recipients (0.9%, 2.5%, and 3.2%, respectively). Correspondingly, continuous public insurance (HR 1.29, 95% CI 1.06-1.56; *p*=0.011), private to public insurance transitions (HR 1.32, 95% CI 1.03-1.71; *p*=0.031), and multiple insurance transitions (HR 1.09, 95% CI 1.07-1.55; *p*=0.020) were associated with increased graft failure risk (**Table S2**). Conversely, transitioning from public to private insurance significantly decreased the risk of graft failure (HR 0.73, 95% CI 0.45-0.92; p=0.019). Additional factors associated with increased graft failure included residence in distressed communities (HR 1.31, 95% CI 1.20-1.41; *p*=0.001) and recipient smoking status (HR 1.24, 95% CI 1.05-1.47; *p*=0.010). Recipients’ college completion (HR 0.81, 95% CI 0.50-1.29; *p*=0.369) was not associated with improved graft survival.

Secondary outcomes also varied significantly by insurance group. Continuous public insurance recipients had the longest average hospital stays (28.01 days; *p*<0.0001) and the highest rates of dialysis requirement post-transplant (14.7%; *p*<0.0001), acute rejection episodes within one year (9.8%; *p*=0.009), and treatments for rejection within the first year (16.6%; *p*<0.0001). Rates of stroke (*p*=0.564) and pacemaker implantation (*p*=0.650) did not significantly differ across insurance trajectories. Recipients with continuous public insurance, transition from private to public insurance, and multiple insurance transitions had significantly higher proportions of moderate-to-severe functional limitations post-transplant compared to those with continuous private insurance and transition from public to private insurance (88.6%, 83.0%, and 75.1% vs. 76.3% and 71.1%, respectively; *p*<0.0001).

## Discussion

In this national study of adult HT recipients in the post-ACA and post-2018 heart allocation policy change, we found that insurance coverage trajectories are strongly associated with post-transplant outcomes. Patients with continuous private insurance had the most favorable outcomes, while those with continuous public insurance experienced significantly worse long-term survival and higher complication rates, including graft failure and acute rejection. Notably, recipients with multiple insurance transitions in the post-transplant period had outcomes nearly as poor as the continuous-public group. HT recipients who switched from private to public insurance had a higher hazard of graft failure and more moderate-to-severe functional limitations, although mortality did not significantly differ from privately insured patients. Collectively, these findings underscore the need for targeted interventions and policies that protect HT patients from insurance instability and its downstream effects.

Our study builds upon and expands on prior research linking insurance status and transplant outcomes. Several studies have shown that HT recipients with public insurance experience worse clinical outcomes on average than those with private coverage. One large registry study found that 5-year survival among Medicaid-insured HT patients was similar to Medicare patients (0.9% lower) but 5.9% lower than among those with private insurance^3^.

Medicaid coverage was also associated with a 77% higher mortality risk beyond two years post-transplant compared to private insurance^3^. Similarly, a recent study of 37,000 HT recipients found that Medicaid patients had an 8% higher hazard of developing cardiac allograft vasculopathy (CAV) within 5 years, as well as 31% higher all-cause mortality and 29% higher graft failure-related mortality^13^. These disparities likely reflect barriers faced by publicly insured patients, including restricted for costly access to immunosuppressive medications, reduced access to specialized care, and broader socioeconomic barriers that delay diagnosis and treatment^1^.

In our cohort, continuous public insurance was associated with a 27% higher adjusted hazard of mortality compared to continuous private insurance, aligning with numerous earlier studies. A prior analysis of UNOS registry data found that Medicare and Medicaid patients had a 18% and 33% higher 10-year mortality risk, respectively, when compared to privately insured patients^14^. Another study reported a 36% higher 1-year mortality hazard for continuous public coverage^8^. Additionally, our study findings suggest that this survival disparity may widen over time: continuous public insurance had increasingly higher mortality hazard at the 1-, 3-, and 5-year timepoints (4.6%, 12.4%, and 15.1%, respectively), relative to continuous private insurance. Similarly, DuBay et al. found that for HT recipients, the mortality hazard for Medicaid patients increased from 1.1 in the first two years post-transplant to 1.8 beyond two years, compared to privately insured patients^3^. These findings reinforce that insurance type is a critical determinant of long-term survival post-HT, with continuous public insurance consistently associated with higher mortality.

Regarding insurance churn, our study found that a single switch from public to private insurance did not significantly affect mortality in adjusted models (HR 1.19, *p*=0.092), nor did a single switch from private to public insurance (HR 1.04, *p*=0.607). In contrast, Tumin et al. in a 2006–2013 pre-ACA cohort, reported a 22% lower mortality hazard for patients transitioning from public to private insurance and a 25% higher hazard for those moving from private to public coverage^8^. A likely reason is that recent policy changes may have reduced the adverse effects of losing private coverage. Medicaid expansion under the ACA increased coverage for low-income adults, enabling more patients with advanced heart failure to be evaluated and listed for transplant while reducing delays in care^1,2,15^. By minimizing cost sharing for critical post-transplant medications (e.g., immunosuppressants) and supporting stable long-term follow-up, it also likely improved continuity of care and graft survival among those transitioning to public insurance^1,2,15^. Notably, a previous study found that from 2000–2020, the share of HT among publicly insured patients increased from 36.7% and 53.4%, with most of the growth after the ACA’s implementation^15^. Another key finding was the effect of multiple-transition insurance trajectories.

Approximately 6% of our cohort experienced more than one insurance change in the first post-transplant year. Their unadjusted 5-year mortality hazard (14.9%) was nearly equivalent to that of the continuous-public group (15.5%) and significantly higher than continuous-private patients (11.9%). After adjustment, undergoing multiple transitions conferred a 19% higher mortality hazard than continuous private insurance. These findings suggest that beyond coverage type, the instability of frequent insurance changes could potentially be detrimental. Churn can introduce administrative hurdles, gaps in coverage for health services or medications, provider network constraints, and overall care fragmentation, all of which could influence post-transplant outcomes^6^. Prior evidence among low-income adults in the general U.S. population shows that insurance churn was associated with lower medication adherence (44.3% vs. 26.3%) and higher emergency department use (18.5% vs. 11.5%) compared to those with stable insurance coverage^6^. Even brief interruptions experienced by HT patients could lead to lapses in immunosuppressive therapy or delayed follow-up, precipitating serious complications^16^. Taken together, our study findings provide further evidence that minimizing insurance instability should be a priority for transplant programs and policymakers.

Our analysis of post-transplant complications further underscores the disparities linked to insurance status. Patients with continuous public insurance had the highest rates of acute rejection, consistent with recent findings by Sakowitz et al., who identified Medicaid insurance as an independent risk factor for CAV, a leading cause of late graft loss^13^. Similarly, adverse outcomes were concentrated in the continuous public insurance subgroup, including longer hospital stays, more acute kidney injury requiring dialysis, and higher rates of moderate-to-severe functional limitations. These patterns suggest that publicly insured patients may face more complex postoperative courses, likely reflecting a combination of poorer pre-transplant health and differences in post-transplant care resources. The consistently worse outcomes among Medicaid and Medicare patients reflects broader structural disparities in transplant care^17^, influencing not only mortality but also a range of post-transplant complications.

Beyond insurance, we also found that lower educational attainment and residence in socioeconomically distressed communities independently predicted higher risks of mortality and graft failure. These findings align with prior research in HT recipients showing that less-than-college level education, Medicare, and Medicaid coverage were each independent predictors of elevated mortality risk −11%, 18%, and 33% higher, respectively, over 10-year mortality risk^14^. Chen et al. similarly found that patients from socioeconomically distressed areas had slightly but significantly lower 5-year survival post-HT (75.3% vs 79.5%), with high-distress areas with a 13% higher mortality hazard^18^. These structural disparities likely reflect differences in health literacy, follow-up access, caregiver support, and neighborhood resources^17,18^. Altogether, these findings suggest that improving HT outcomes will require not only promoting insurance stability but also addressing the upstream social determinants that place socioeconomically vulnerable patients at greater risk.

### Policy Implications

These findings have several important policy implications. First, given the association between public insurance, insurance churn, and poorer outcomes, ensuring stable coverage and adequate financial support across the HT care continuum should be a priority. This might include extending transitional insurance support or disability-based coverage beyond the current limits. Many HT patients qualify for Medicare due to heart failure but risk losing it if they recover and no longer meet disability criteria. By 2020, nearly half of HTs were funded by public insurance, giving the Centers for Medicare & Medicaid Services (CMS) leverage to standardize care and negotiate broader, more consistent coverage^15^. For example, the model of lifetime Medicare coverage for kidney transplant immunosuppressants could be adapted to HT, ensuring patients do not lose coverage abruptly^19^. Strengthening the safety net during unavoidable transitions (such as aging out of a family insurance plan, job loss, or changes in Medicaid eligibility thresholds) could further prevent critical treatment disruptions. Strategies could include leveraging case management to facilitate prompt re-enrollment, and policies requiring grace periods or temporary coverage extensions so that access to essential procedures and medications are not interrupted during administrative turnover.

Second, improving quality of public insurance is essential. Expanding coverage alone is insufficient if it does not translate into equitable, effective care. Medicaid reimbursement rates are substantially lower than those of private insurance (often covering only 58 to 65 cents on the dollar), which can limit transplant centers’ ability to absorb the intensive costs of managing complex HT patients^20^. Additionally, public insurance plans tend to have narrower provider networks, further restricting access to care. In a 2019 national analysis of cardiology networks, Medicaid managed-care plans included on average just 46% of nearby cardiologists compared with 69% in large-group employer networks^21^. Another report found that Medicaid plan networks included over 60% fewer cardiologists than commercial plans in certain cities^22^. Public insurance has also been linked to higher cost-sharing for essential post-transplant medications.

For example, Medicare’s standard 20% copayment on immunosuppressants places a substantial burden on many transplant recipients, potentially increasing nonadherence and compromising graft survival^2^. Achieving more equitable outcomes thus requires strengthening coverage itself: raising reimbursement rates or offering transplant-specific supplemental benefits, enforcing minimum network standards for cardiology and transplant subspecialties, and reducing out-of-pocket costs for post-transplant medications and follow-up care. Further, addressing upstream barriers by funding wrap-around services—such as transplant pharmacists, transportation support, and telehealth—and expanding these through CMS quality initiatives could also help close the gap in the accessibility and affordability of care between public and private plans.

### Limitations

This study has several limitations. First, it relied on national transplant registry data (UNOS/OPTN), which depends on accurate reporting by transplant centers. Insurance status was recorded at only three timepoints (listing, transplant, and 1-year follow-up), thus, more frequent changes or brief coverage gaps may not have been captured, potentially leading to misclassification. Second, our broad classification of insurance into “private,” “public,” or “other” masks meaningful heterogeneity. We combined Medicare and Medicaid as “public insurance” to ensure statistical power and simplify trajectories, although these groups differ considerably;

Medicare recipients are typically older or disabled, while Medicaid patients are often younger and lower income, with distinct health profiles and risks. Similarly, “private insurance” includes a wide range of plans with varying benefits and provider networks. We also did not separately analyze dual-eligible patients because of limited statistical power. This is an important distinction as prior studies have shown that dual eligible patients with heart failure or acute myocardial infarction have more baseline comorbidities, greater healthcare utilization, longer hospital stays, higher 30-day readmission rates, and increased short-term mortality^23,24^.

Third, given the observational study design, residual confounding remains a concern despite adjustment for demographic, clinical, and socioeconomic factors. Unmeasured variables such as frailty, cognitive impairment, or caregiver availability may have influenced both insurance transitions and outcomes. We also lacked detailed data on key mediators like immunosuppressant adherence, drug monitoring, and biopsy protocols, which could affect the associations between insurance type and post-transplant outcomes. Reverse causation is also possible; for example, health deterioration might lead to employment loss and a switch to Medicaid, which could overestimate the hazard of post-transplant mortality or complications in the public insurance subgroups. Fourth, our outcome measures have inherent limitations. For example, the definition of graft failure, based on ISHLT criteria, could be subject to misclassification, and we did not explore more specific causes such as CAV that other studies have examined^13^. Finally, because this study was limited to the United States, our findings are not generalizable to other countries with different health insurance structures and rates of churn. Even within the U.S., substantial variability exists across state Medicaid programs, thus, our national analysis may have obscured important state-level differences.

## Conclusion

This national cohort study shows that insurance patterns strongly influence post-transplant outcomes. Continuous private insurance confers the best survival and lowest complication rates, while continuous public coverage and frequent insurance changes are linked to higher risks of rejection, graft failure, and mortality. Insurance churn may disrupt continuity of care, compounding risks for socioeconomically disadvantaged patients. These findings underscore the need for policies and transplant programs that secure stable, adequate insurance coverage to ensure equitable benefits of heart transplantation for all patients.

## Data Availability

The data that support the findings of this study are available from the Organ Procurement and Transplantation Network (OPTN) and the United Network for Organ Sharing (UNOS) Standard Transplant Analysis and Research (STAR) files. Restrictions apply to the availability of these data, which were used under license for this study. Researchers may request access to the STAR files through the OPTN/UNOS data request process (https://optn.transplant.hrsa.gov/data/request-data/). No additional, unpublished data are available.

## Acknowledgements

We are also grateful to the Pauley Heart Center for providing resources and research support.

## Sources of Funding

None

## Disclosures

None

## Abbreviations

ACA: Affordable Care Act
BMI: Body Mass Index
CAV: Cardiac Allograft Vasculopathy
CMS: Centers for Medicare & Medicaid Services
DCI: Distressed Communities Index
DCD: Donation after Circulatory Death
DPP: Direct Procurement and Preservation
ECMO: Extracorporeal Membrane Oxygenation
HR: Hazard Ratio
HT: Heart Transplantation / Heart Transplant
ISHLT: International Society for Heart & Lung Transplantation
LOS: Length of Stay
Medicaid: U.S. Public Health Insurance Program for low-income individuals
Medicare: U.S. Public Health Insurance Program for elderly and disabled
NRP: Normothermic Regional Perfusion
OPTN: Organ Procurement and Transplantation Network
SD: Standard Deviation
STAR: Standard Transplant Analysis and Research
UNOS: United Network for Organ Sharing
VA: Veterans Affairs
VIF: Variance Inflation Factor

## Notes

### Competing Interest Statement

The authors have declared no competing interest.

### Clinical Trial

N/A

### Author Declarations

This study was reviewed by the Virginia Commonwealth University Institutional Review Board and determined to be exempt from review because it used de-identified, publicly available data from the Organ Procurement and Transplantation Network/United Network for Organ Sharing Standard Transplant Analysis and Research files. All data use complied with relevant federal and institutional guidelines and regulations.

